# Spatiotemporal Correlation between Obesity Prevalence and the Percentage of Households using Air-conditioners in the United States

**DOI:** 10.1101/2022.10.12.22281015

**Authors:** Evan Li, Alexander Wang, Carter Xu, Felix Twum, Jian Zhang

## Abstract

**Purpose:** The axiom that obesity epidemic was driven by steady increases in energy intake has been challenged by empirical evidence, including the failure to make meaningful progress either treating or preventing obesity. Adopting new paradigm were urgently needed. We aimed to collect ecological evidences to support an alternative hypotheses that increased usage of air-conditioners (AC) may contribute to obesity epidemic in the U.S.

**Method:** U.S. national survey data were retrieved from public domains, including the % of homes with AC units (AC ownership), dietary energy intake, and obesity prevalence. Correlation efficient were estimated, and Joint point regressions were ran to describe time segments and estimate annual percentage change (APC).

**Results:** Nationally, Obesity prevalence significantly associated with the increasing trend of AC ownership (partial *r*=0.046, and *p* < 0.05) not dietary energy intake (r < 0.0001, and *p* = 0.58). When stratified by region, the rank of obesity prevalence across regions was consistent with that of AC ownership throughout the study period; the South led the increasing trends for both AC ownership and obesity prevalence. The climbing national trend of obesity slowed down around 2002 (before 2002 APC=4.38, and APC=2.22 after) following the saturation of AC penetration in the South (before 1994, APC=1.04, *p* < 0.05, and APC=0.05 after 1994, *p* > 0.05.

**Conclusions:** Spatiotemporal correlations support the hypothesis that penetration of AC may contribute to the obesity epidemic. Further investigation may lead to novel therapies and a new avenue to explore the strategies addressing twin clusters of pandemics, obesity, and climate change.

## 1. Introduction

Considered axiomatic, the commonly accepted explanation for the increase in the prevalence of obesity - pervasive overeating, has been increasingly challenged [1, 2], largely due to the failure to make meaningful progress in either treating or preventing obesity [3] [4]. Rigorously collected national data suggest that over the last two decades, Americans have been eating relatively less for the larger body size while the climbing trend of obesity continues without reversing signs in view. The discordance between theory and evidence calls for a new paradigm of obesity research [2, 4, 5], most recently amid the covid-19 pandemic [1, 6].

One ubiquitous factor influencing both energy expenditure and energy intake is the ambient or environmental temperature in which we live [7]. Along with elevated living standards and advancements in technology, air-conditioners (AC) have been increasing significantly and ambient temperatures have homogenized, significantly buffering humans from extreme temperatures and decreasing human beings’ exposure to mild seasonal variations of ambient temperature. Humans today spend an increasing amount of time in a thermally tight-controlled state. After systematically reviewing the evidence from animal studies, human experiments, and epidemiological studies, McAllister and her colleagues hypothesized that increased usage of ACs and more time spent in the thermal neutral zone may result in a positive energy balance and manifest as weight gain in a person, and increasing obesity prevalence at the population level [2].

Following McAllister et al’s preliminary hypothesis, mounting biological evidence was collected by Mavrogianni et al. [5], Moellering and Smith [7] to support the causal relationship between tightly controlled ambient temperature and the development of obesity. However, due to methodological challenges, epidemiological evidence showing the effects of variation in a long-term thermal environment on energy balance or body weight remains scarce. The direct epidemiological evidence is still limited to the ecological figure correlating the percentage of homes with air conditioning with the obesity trend for the period between 1978 and 1998 in the United States presented in McAllister et al.’s hypothesis-generating report published more than 10 years ago. With additional years included, and spatial dimension incorporated, we performed this ecological assessment to examine the spatiotemporal correlation between obesity prevalence and the percentage of households using ACs in the United States where both obesity prevalence and AC utilization have been soaring in the last several decades.

## 2. Materials and Methods

### 2.1. Data source and variable definitions

We retrieved the series of data on obesity prevalence, population means of dietary energy intake, and the percentage of homes with AC units, from well-established continuous national surveys, starting from 1970.

#### 2.1.1. The percentage of homes with AC units (AC ownership)

The Survey of Construction (SOC) has been conducted by the U.S. Census Bureau to obtain current national and regional characteristics of new, privately-owned single-family and multifamily housing units [8]. Starting in 1959, and partially funded by the Department of Housing and Urban Development, SOC has collected the physical characteristics of each housing unit, such as square footage and whether AC is installed. A multi-stage stratified random sample procedure was used to provide nationwide coverage. Approximately, 900 building permit-issuing offices and more than 70 land areas not covered by building permits were selected. A trained field representative contacts the owner or builder selected by telephone or in person, to conduct the interview each month. Estimates are adjusted to reflect variations by region and type of construction, data are seasonally adjusted for late reports, and houses started or sold before a permit was issued. The data on AC ownership were available for the years between 1973 and 2021.

The data collected by SOC has been used by the Bureau of Economic Analysis for the development of the national income and product accounts, The Federal Reserve Board and Council of Economic Advisers to determine the condition of the economy, and the Department of Housing and Urban Development to develop and evaluate housing programs. The data have also been widely used by business communities to plan production schedules and establish market shares, adjust rates and establish replacement costs, and estimate mortgage demand [9]. We selected the presence of AC in new single-family houses completed as the representative estimates for AC ownership in current analyses. SOC data are released at the national level and by Census Region. However, the Census Region has been defined from geography and history instead of the climate (Supplemental **Figure 1**), making the homogeneity of AC ownership region specific.

#### 2.1.2. Data on obesity prevalence and definition of obesity

The data on obesity prevalence were from two major national sources, The National Health and Nutrition Examination Survey (NHANES) and the Behavioral Risk Factor Surveillance System (BRFSS), both conducted by the Centers for Disease Control and Prevention (CDC).

NHANES is a continuous survey designed to assess the health and nutritional status of adults and children in the United States. Combining interviews and physical examinations, NHANES is a major program to produce vital and health statistics for the Nation. Authorized by the National Health Survey Act passed in 1956 and managed by the National Center for Health Statistics of CDC, the NHANES began in the early 1960s and had been conducted as a series of surveys with 10 years apart. In 1999, the survey became continuous, and the data have been released biannually. The survey examines a nationally representative sample of about 5,000 persons each year. The examination component consists of medical, dental, and physiological measurements administered by highly trained medical personnel. Findings from NHANES have been used to determine the national prevalence of major diseases and risk factors for diseases, served as the basis for national standards for such measurements as height, weight, and blood pressure, and help develop sound public health policy, direct and design health programs, and expand the health knowledge for the action [10]. The prevalence of obesity in children and adolescents estimated by Fryar et al[11], and among emerging adulthood estimated by Ellison-Barnes et al [12] using NHANES data were correlated with AC ownership in the current report. The data on the national obesity prevalence were available for the years between 1977 and 2015.

The BRFSS is the nation’s premier telephone survey system to collect state data about U.S. residents regarding their health-related risk behaviors and chronic health conditions. Starting in 1984 with 15 states, BRFSS currently covers all 50 states as well as the District of Columbia and three U.S. territories. It uses a multistage cluster design based on random digit dialing methods of sampling to select a representative sample from each state’s noninstitutionalized civilian residents aged 18 years or older. By collecting data at the state and local levels, BRFSS has been developed into a powerful tool for building health promotion activities. Every year, more than 400,000 adults are interviewed, making BRFSS the largest continuous health survey system in the world. A detailed description of the survey methods has been published elsewhere [13]. Each year, state health departments use the procedures standardized by BRFSS to collect data through a series of telephone interviews with U.S. adults. Obesity was defined as a body mass index (BMI) of 30 or higher, and BMI was a measure of an adult’s weight in relation to his or her height, specifically the adult’s weight in kilograms divided by the square of his or her height in meters. Height and weight data are self-reported in BRFSS. Region-specific obesity prevalence was obtained by combining population-weighted state prevalence. The data on obesity prevalence by the state were available for the years between 1985 and 2020.

#### 2.1.3. Dietary energy intake

Nutrition assessment is one of the key topics of the NHANES. A 24-hour dietary recall interview was administrated by trained dietary interviewers (mostly dietitians) through an automated interview using the Dietary Data Collection (DDC) system. Special probes were used to help recall commonly forgotten items such as condiments, accompaniments, fast foods, alcoholic beverages, etc. It provided such information as specific food items and their quantities ingested for all regular meals, foods, or snacks consumed between meals on the day, midnight to midnight, preceding the interview. The types and amounts of foods consumed were recalled using recall aids such as special charts, measuring cups, and rulers to help quantify the amounts consumed. Each food item was assigned a unique food code, and the approximate portion was coded. The food codes matched the nutrient composition of over 3000 food items obtained from the US Department of Agriculture, food manufacturers, and other sources. From the above information, all food ingested during the 24-h period was then reduced by a computer program to standard units of measure for actual dietary intake during the 24-hour recall period. The data were then linked to the US Department of Agriculture’s Survey Nutrition Database and produced total energy intake [14]. The dietary energy intake among adults between 1976 and 2010 was estimated by Ford and Dietz [15], and the intake between 1999 and 2018 was estimated by Mozarffarian [1]. The difference between the two reports across the overlapped period (1999-2010) was used to adjust the continuity. Hence, the data on the national average of dietary energy intake were available for the years between 1976 and 2018.

### 2.2. Analytic steps

We used SAS (SAS Institute, Cary, NC, SAS 9.4) and Joinpoint Trend Analysis Software (National Cancer Institute, Bethesda, MD, Joinpoint V4.9.0.0) to conduct analyses. A reliable assessment of obesity prevalence in adults and dietary energy intake was not available for sub-national units. The ecological correlations were limited to the secular trend without spatial components at the national level. For visual assessment, we used PROC REGRESSIONPLOT of SAS with a degree of 3 to draw trend curves for AC ownership, dietary energy intake, and obesity prevalence by age group, and the join points within each curve were identified with Joinpoint Trend Analysis Software[16], and annual percentage changes (APCs) of data series were estimated for each time segment identified. Mixed models with repeated measures were used to adjust for autocorrelation and the coefficients of partial correlation between AC ownership, dietary energy intake, and obesity prevalence across the study period were calculated using the formula proposed by Lipsitz et al [17].

The obesity prevalence by region was estimated by combining state prevalence weighted by state populations. Since no region-specific dietary energy data were available, no effort was made to estimate the partial correlation when the assessment was stratified by region. The increase in obesity prevalence (%) corresponding to a unit increase of AC ownership (%) was estimated by region with linear regression without autocorrelation adjustment.

## 3. Results

The contiguous United States has a broad range of climates between regions (Supplemental **Figure 2**). Climate variates also significantly within the region. Spanning across 20 degrees of latitude, with landlocked desert communities and mild temperatures communities along the coastline and two noncontiguous states (Alaska and Hawaii), the vast geographic differences across the West make the homogeneity of AC penetration statistics in the West less comparable to other regions, which span about 10 degrees of latitude. AC penetration increased at different rates across regions (**Table 1**). The inclusion of air conditioning in homes was highest in the South. By 1985, more than 90% of homes had ACs present, and almost 100% have ACs installed in the South by 2020. In contrast, less than 50% of homes in the Northeast had AC in 1985 and still less than 90% of the home have installed an AC unit in 2021. Similarly, adult obesity also showed a large variation at the beginning of the study period and has been converging toward the end of the study period. The obesity prevalence was twice as high in Midwest and South than that in the Northeast and West. At the end of the study period, all regions had a mean obesity prevalence greater than 30%. It’s also noted that the population in the South accounts for more than one-third of the national total, and the proportion increased from 0.34 in 1990 to 0.38 in 2020.

**Table 1.**
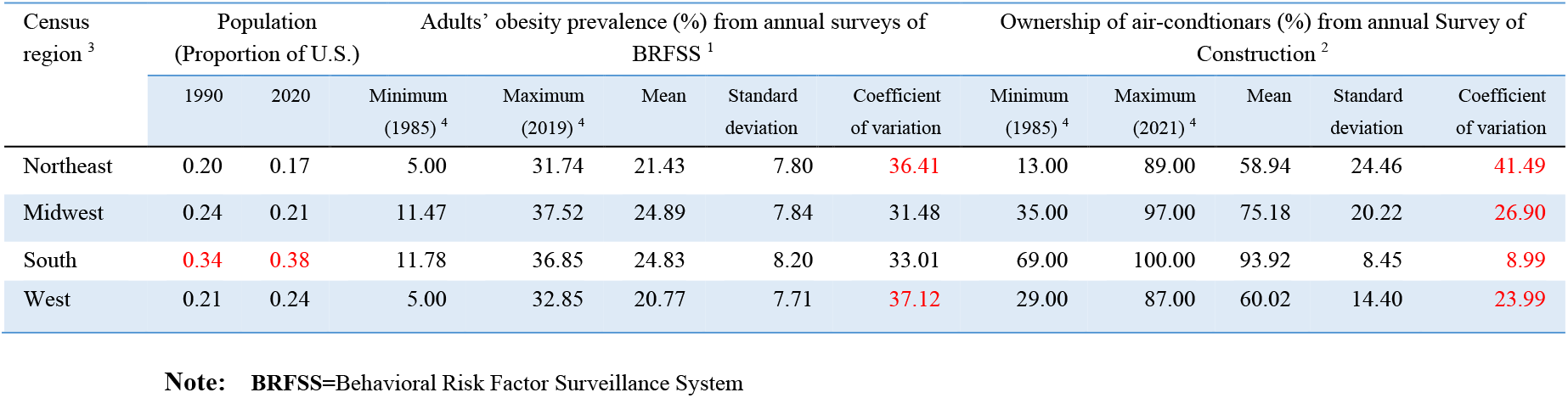
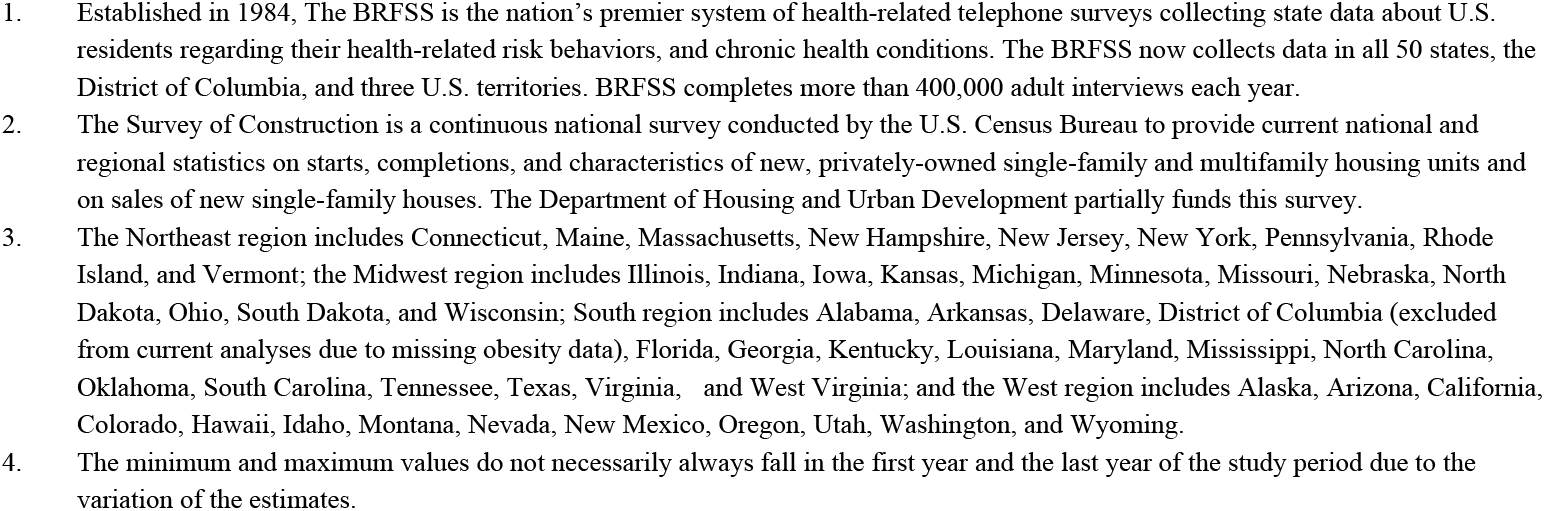
The Basic Characteristics of 4 Census Regions, United States, 1985 – 2019

| Census region <sup>3</sup> | Population (Proportion of U.S.) |  | Adults' obesity prevalence (%) from annual surveys of BRFSS <sup>1</sup> |  |  |  |  | Ownership of air-conditioners (%) from annual Survey of Construction <sup>2</sup> |  |  |  |  |
| --- | --- | --- | --- | --- | --- | --- | --- | --- | --- | --- | --- | --- |
|  | 1990 | 2020 | Minimum (1985) <sup>4</sup> | Maximum (2019) <sup>4</sup> | Mean | Standard deviation | Coefficient of variation | Minimum (1985) <sup>4</sup> | Maximum (2021) <sup>4</sup> | Mean | Standard deviation | Coefficient of variation |
| Northeast | 0.20 | 0.17 | 5.00 | 31.74 | 21.43 | 7.80 | 36.41 | 13.00 | 89.00 | 58.94 | 24.46 | 41.49 |
| Midwest | 0.24 | 0.21 | 11.47 | 37.52 | 24.89 | 7.84 | 31.48 | 35.00 | 97.00 | 75.18 | 20.22 | 26.90 |
| South | 0.34 | 0.38 | 11.78 | 36.85 | 24.83 | 8.20 | 33.01 | 69.00 | 100.00 | 93.92 | 8.45 | 8.99 |
| West | 0.21 | 0.24 | 5.00 | 32.85 | 20.77 | 7.71 | 37.12 | 29.00 | 87.00 | 60.02 | 14.40 | 23.99 |

Note: BRFSS=Behavioral Risk Factor Surveillance System

Nationally, no ecological correlation was observed between dietary energy intake and obesity. Actually, while obesity continuously increased during the study period, dietary energy intake decreased starting from 1999 at an APC of 0.25 (***p*** < 0.05, **Figure 1**). AC ownership significantly correlated with obesity prevalence, especially in older adolescents and emerging adults (Supplemental **Table 1**). The adjusted partial correlation coefficient was less than 0.001 for the correlation between obesity prevalence and dietary energy intake, and 0.046 (***p*** < 0.001) for the correlation between obesity and AC ownership. AC ownership increased significantly throughout the study period with an APC of 0.74 (***p*** < 0.001), and obesity prevalence also increased significantly but slowed down starting from 1999. The APC for emerging adults (aged 18-25 years) was 6.48 (***p*** < 0.05) for 1978 – 1999, and 0.74 (***p*** < 0.05) for 1999 - 2015 (the curve in magenta color).

**Figure 1:**
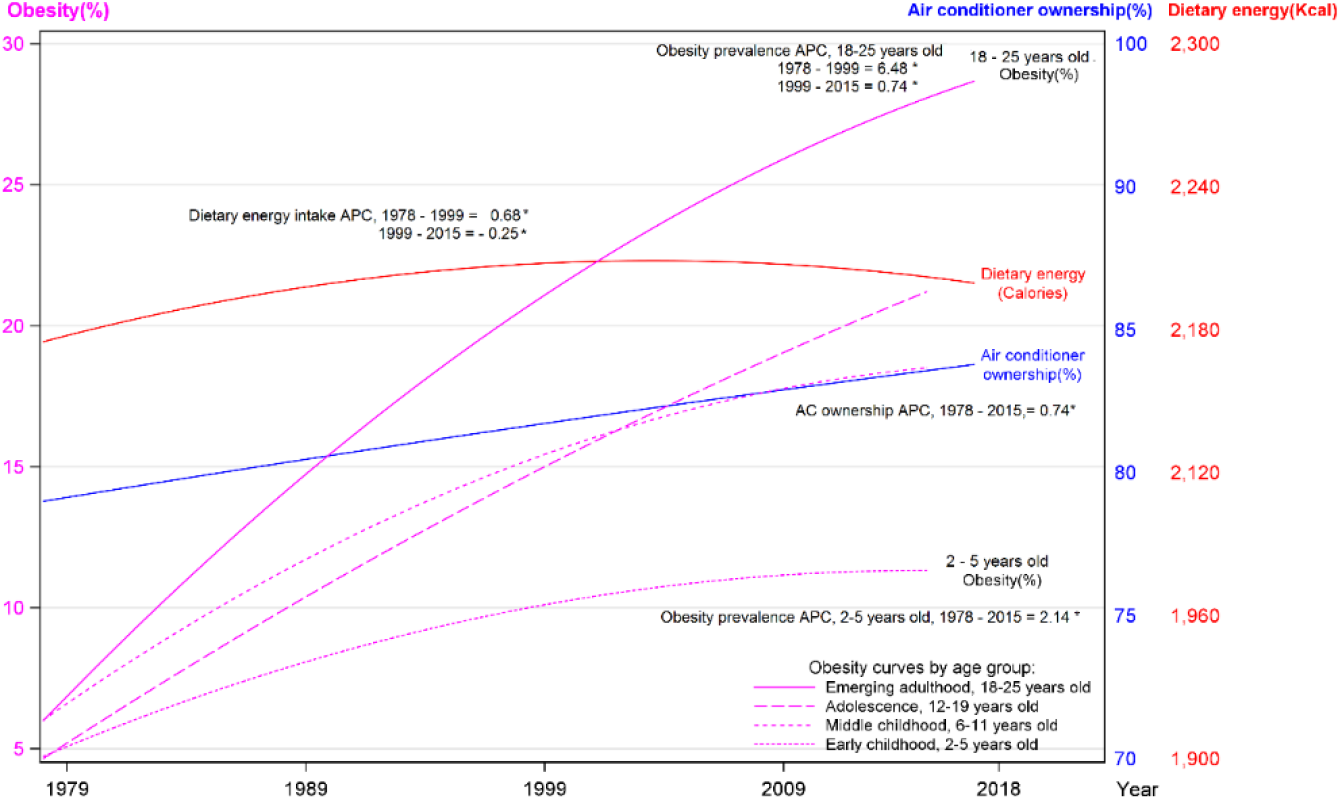
The National Trend of Pediatric Obesity, % of Homes with AC units 1977 – 2020, the United States ^1,2,3,4^ 1. APC = Annual percentage change, * indicates that the APC significantly different from zero at the alpha = 0.05 level. 2. Data source: The data on AC utilization were collected by Annual Characteristics of New Housing Surveys conducted by the US Census Bureau. 3. For childhood obesity, the measured weight and height were translated into sex- and age-specific BMI percentiles of 2000 CDC Growth Charts, and children were categorized as obese if they had sex- age-specific BMI ≥ the 95th percentile. The estimations were made by Fryar et al.11 For obesity in emerging adulthood, the estimations were made by Ellison-Barnes et al.12 4. The adults’ dietary energy intakes were based on 24-hour dietary recall data from NHANES. For the year 1977 – 2010, data from adults aged 20–74 y were analyzed, and the means were adjusted for age, sex, race or ethnicity, educational status, and BMI, where applicable as detailed by Ford and Dietz. 15 For the year 2011-2018, the estimates were made by Mozaffarian 1 without specifying whether the estimates were crude or adjusted. The mean of the estimate difference (47.3 cal) for the overlap of the period (1999-2010) was used to adjust the estimate from the two reports for continuity.

When stratified by region, the rank of obesity prevalence by region was consistent with that of AC ownership roughly throughout the entire study period (**Figure 2**), the South (in black color) and Midwest region (red color) led the increasing trends for both AC ownership and obesity prevalence. The AC penetration reached the saturation point around 1994 in the South (APC=1.04, p< 0.05 before 1994, and APC=0.05 after 1994, p > 0.05, data not shown), and the increasing trend of obesity at the national level apparently was slowing down following the saturation of AC penetration in the South, and slowing down of obesity in the South (before 2002 APC=4.38, and APC=2.22 after). It was also noticed that the increasing trend of AC ownership slowed down slightly between 1994 and 2000 in other regions. Although both AC ownership and obesity prevalence had already been at the highest level in the South among four regions at the beginning of the study period, the correlation remains stronger in the South and Midwest compared to that in the Northeast region during the study period, the increase of obesity prevalence in adults associated with one percent increase of AC ownership were 1.45%, 0.76%, and 0.54% for South, Midwest, and Northeast region respectively (**Figure 3**).

**Figure 2:**
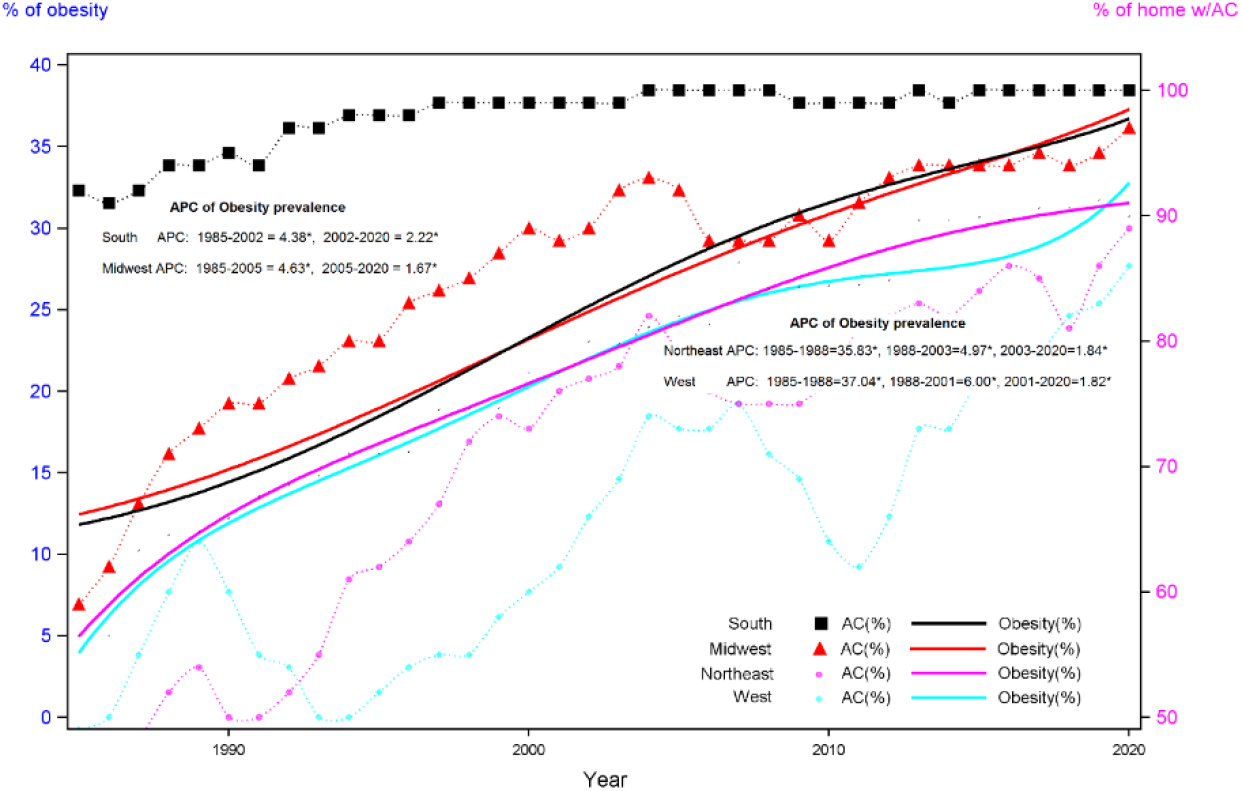
The Obesity Epidemic and Trend of AC ownership Led by South and Midwest Regions, U.S. 1985 – 2020 1. APC = Annual percentage change, * indicates that the APC significantly different from zero at the alpha = 0.05 level. 2. The BMI data in this figure were collected through CDC’s Behavioral Risk Factor Surveillance System. The data on AC utilization were collected by Annual Characteristics of New Housing Surveys conducted by the US Census Bureau. 3. Body Mass Index (BMI): A measure of an adult’s weight in relation to his or her height, specifically the adult’s weight in kilograms divided by the square of his or her height in meters. Obesity was defined as a BMI of 30 or higher.

**Figure 3:**
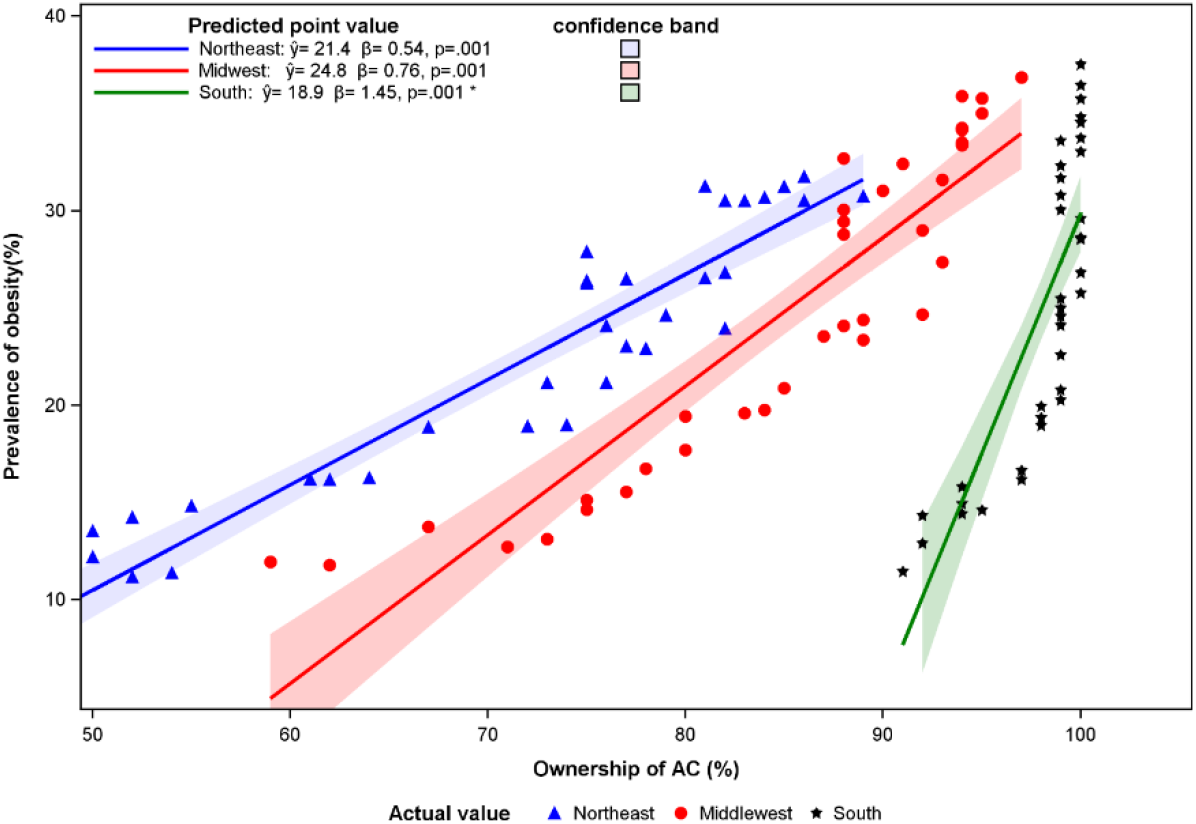
Correlation between adult obesity prevalence and air-conditioner ownership, South, Northeast, and Midwest Regions, 1985 – 2020, United States * The estimates for the South were from the regression on the data ending at 2005 when the AC ownership reach 100%.

Further examination by states indicated that West Virginia and Mississippi consistently led the increasing trend of obesity (**Table 2**).

**Table 2:**
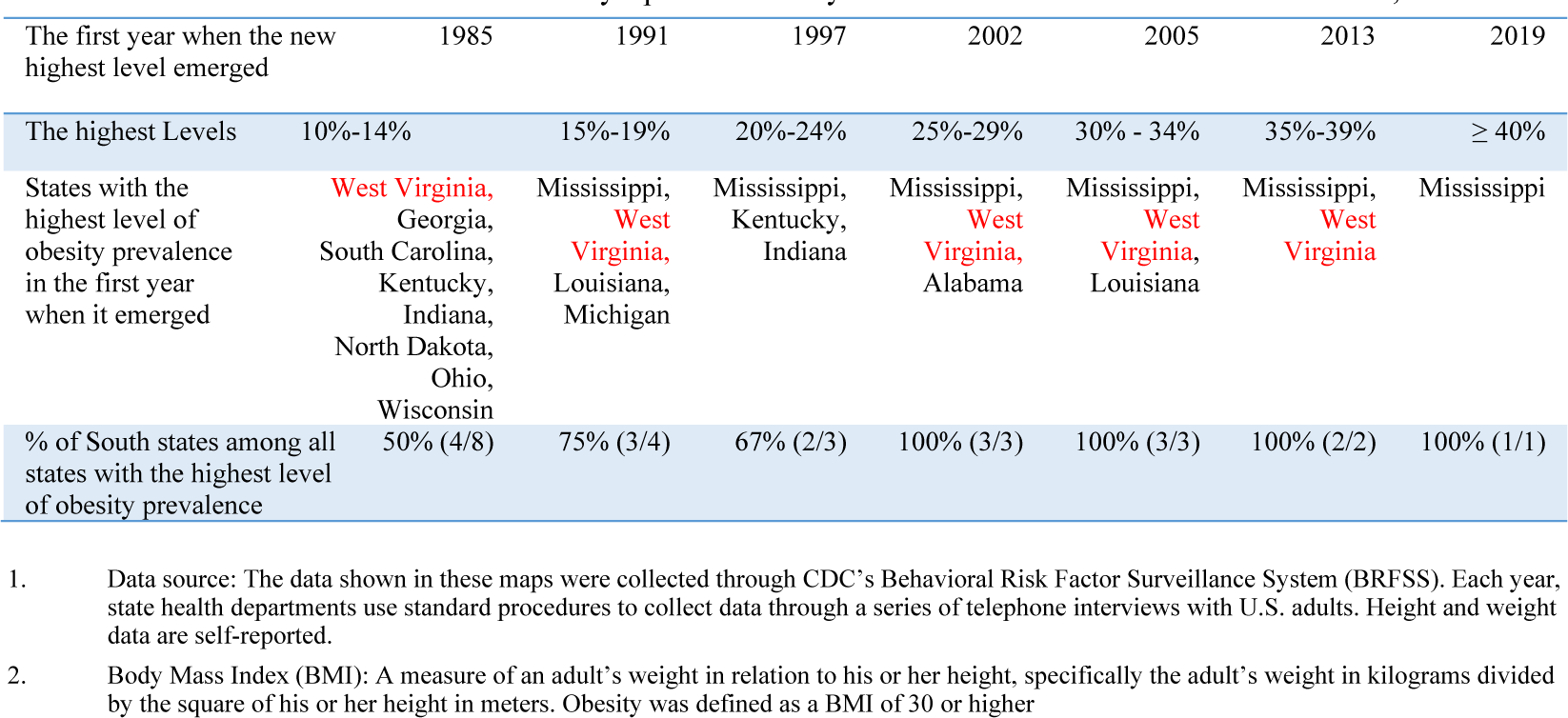
The Obesity Epidemic Led by the States in the South in the United States, 1985 – 2019

## 4. Discussion

The evidence compiled in the current report from spatiotemporal correlations between obesity prevalence and the percentage of homes using ACs supports the hypothesis that penetration of air-conditioning may contribute to the obesity epidemic. AC ownership increased throughout the study period, and obesity prevalence also increased significantly but slowed down around 1999 – 2000 after AC penetration reached the saturation point in the South, the region leading the obesity epidemic in the United States. The correlation between obesity prevalence and AC ownership was region-specific; Obesity prevalence increased more rapidly in the region with high AC penetration, and the obesity prevalence was relatively low in the regions where AC penetration was less saturated.

One of the most puzzling observations in today’s obesity-related research is the discordance between dietary energy intake and the national obesity trend [1, 6]. The dietary energy intake among U.S. adults started to decline since the 1999-2000 national survey, while the nation became more obese. The finding from the current analysis offers a new explanation for this discordance. While obesity continuously increased during the study period, the increase slowed down starting from 1999, the year when AC penetration reached a saturation point i.e., 100% of the home having AC installed, in the South, the region that led the obesity epidemic and experienced a significant increase in both AC ownership and obesity prevalence. There is a possibility that the slowing down of the national climbing trend of obesity may be due to the lost moment created by AC penetration in the South.

The physiological mechanism underlying the connection between ambient temperature and obesity has been well described by Mavrogianni et al.[5], Moellering and Smith [7]. Briefly, the thermoneutral zone (TNZ) is defined as the temperature range where neither heat generation nor dissipation is required for thermal homeostasis [18]. When the ambient temperature goes below the low limit of TNZ, adaptive and thermogenic mechanisms are activated to combust lipids and glucose, resulting in heat production and core body temperature preservation [19, 20]. When the ambient temperature rises above the upper limit of TNZ, physiologic processes to amplify heat dissipation are initiated, resulting in a metabolic increase [21, 22]. Living either above or below the boundaries of TNZ increased energy expenditure [21-23]. As humans spend more time in the thermal comfort zone, metabolic demand at thermoneutrality has declined [23]. Meanwhile, dietary energy intake kept climbing in the latter half of the last century, and the resultant energy imbalance has driven obesity prevalence to soar in the last three decades of the 20th century. Dietary energy imbalance, exacerbated by tightly controlled ambient temperature may contribute more than the putative contributors commonly discussed to the obesity epidemic. Increasing numbers of studies also reported a meaningful interaction between leptin and ambient temperature [24-26]. Increased usage of AC likely results in the downregulation of leptin’s role in maintaining energy homeostasis.

Despite the consistent evidence from animal studies [27] [28] [29], human experiments [30-38], and extensive hypothesis formatting efforts [7], the epidemiological evidence remains limited [5]. With the premise that weight gain during the first four months of life is positively correlated with childhood overweight status [39], and childhood overweight would be carried on toward adulthood [40, 41], McAllister et al collected studies on the relationship between perinatal thermal environment and the risk of obesity as the indirect epidemiological evidence to support the causal relation between AC utilization and obesity [42]. The direct epidemiological evidence to support the relationship between tightly controlled ambient temperature and the risk of obesity remains the ecological correlation presented by McAllister et al in their initiative report hypothesizing the causal relationship between AC use and obesity [42]. Our report provides an additional spatial dimension with an extended temporal trend.

The ecological evidence is relatively weak, and not as robust as those from individual-based epidemiologic studies due to the inability to control for potential confounders from ecological data. Opportunities for alternative methods should not be discouraged in the enthusiasm for individual-based epidemiologic studies. It is often impractical or unethical to randomly assign people to be exposed to different air-conditioned living environments with sufficient fidelity on a large scale and for a sufficiently lengthy period to permit confident conclusions about long-term population effects from AC utilization [2]. Thus, alternative research using complementary design as presented in the current report offer additional perspective to an overall body of evidence. With the lacking capacity to control for confounders being a major concern, the leading limitations of the ecological assessment must be acknowledged. It has been well documented that obesity disproportionately affects racial minority groups [43], the spatial data used in the current analysis may be confounded by the proportions of racial minority populations. However, the two states that consistently led the climbing obesity trend was ranked roughly at two ends of the population ethno-racial spectrum, with Mississippi having the highest and West Virginia having a relatively low percentage of minority, indicating the spatiotemporal correlation presented in the current report is unlikely to be confounded by the ethno-racial composition of the population.

It is worth noting that dietary energy intake started to plateau or decline around 1999-2000 [1], the years when AC penetration reached the saturation point in the South. Due to the ecological nature and lack of region-specific dietary data, the current study cannot rule out the possibility that the slowdown of the climbing trend of obesity prevalence might be also explained by the decline of dietary energy intake, in addition to the losing momentum of AC penetration in the South. AC utilization might be desirable over AC ownership to be used to correlate with obesity prevalence since there might be a dose-response association between conditioned ambient temperature and the development of obesity. Behavioral adaptations such as clothing adjustments were not counted. Workplace temperature and temperature-controlled transportation are all parts of the exposure of interest, but not included in current analyses. The census region has been defined based on geography and history instead of the weather. The homogeneity of AC utilization within the region would be poor, and the misclassification of states within each region, especially the AC utilization, would potentially depreciate the correlations investigated. In self-reported studies like BRFSS, People tend to underestimate their weight and overestimate their height, resulting in an underestimation of obesity prevalence [44, 45]. Most relevantly, the underestimation may change over time as the popularity of the body-positivity movement increases [43]. As a phone-based data collection system, BRFSS is not able to sample the people without telephones, and such individuals are likely to be of lower socioeconomic status, a factor well documented to be associated with obesity. The data on obesity from BRFSS is likely a conservative estimate [46]. These limitations taken together would weaken the correlation rather than create a spurious spatiotemporal correlation.

## 5. Conclusions

As global warming has escalated, the penetration of air conditioning would be more aggressive, especially, in the developing world where income is rising, and urbanization is accelerated [47] Comfort cooling represents one of the largest end-use risks to our climate [47], the evaluation of its direct impact on health has been limited to the positive side, such as increasing the productivity of healthy people [48, 49], and the survivability of vulnerable populations [50, 51]. More efforts are needed to examine the impact on overall health. Studies examining the relationship between AC utilization and obesity may lead to the development of novel therapies to address obesity on an individual level and open a new avenue to explore the new strategies to address the twin clusters of pandemics, climate change, and obesity. The spillover effects of these efforts would be profoundly extended to global Sustainable Development Goals, including international trading, economic & financial stability, and global security [52].

## Data Availability

All data produced in the present study are available upon reasonable request to the authors.

https://wwwn.cdc.gov/nchs/nhanes/Default.aspx

https://www.cdc.gov/nchs/data/hestat/obesity_child_15_16/obesity_child_15_16.pf

https://www.cdc.gov/obesity/data/prevalence-maps.html#downloads

https://www.census.gov/construction/chars/

https://www.census.gov/construction/chars/definitions/#r

## Acknowledgments

The NHANES, BRFSS, and Survey of Construction have been developed and funded by multiple federal agencies and operated by the Centers for Disease Control and Prevention, and the U.S. Census Bureau. We thank Yanfeng Li, MD. MPH, Centers for Disease Control and Prevention, for assistance with statistical analyses. No compensation was provided for these consultant services. The views presented in the current report are from neither Yanfeng Li nor her affiliation.

## Conflict of Interest Disclosures (includes financial disclosures)

The project was conducted by E.L., A.W, and C.X. as the 2022 summer activities of the joint research club of Northview High School in Fulton County (Evan Li) and Lambert High School in Forsyth County (A.W, and C.X.). Parts of the analytic effort were conducted as the course exercise of Advanced Placement Statistics. No funding was obtained to conduct the study, and all co-authors declare that they have no conflicts of interest.

## Ethics Approval and Informed Consent Statement

All data used in the current analysis were available from publicly accessible repositories and de-identified. This analysis was considered by the Georgia Southern University (GSU) Institutional Review Board as exempt from approval. The GSU IRB committee also exempted this study from receiving informed consent.

## Author’s Contribution

Conceptualization, J.Z. and F.T.; Methodology, J.Z.; Software, J.Z.; Validation, J.Z., and F.T.; Formal Analysis, J.Z.; Investigation, all authors.; Resources, J.Z.; Data Curation, all authors; Writing – Original Draft Preparation, J.Z..; Writing – Review & Editing, all authors; Visualization, all authors; Supervision, J.Z.; Project Administration, E.L.; Funding Acquisition, not applicable.

## Data Availability Statements

The map is available from at US Census Bureau (https://www.census.gov/construction/chars/definitions/#r) accessed on Sept 10, 2022, and are not protected by copyright in the United States (17USC§ 105). The BMI data of children and adolescents were calculated from CDC’s National Health and Nutrition Examination Survey, downloaded from the Centers for Disease Control and Prevention (https://wwwn.cdc.gov/nchs/nhanes/Default.aspx). The data used in the current report on pediatric obesity were estimated by National Center for Health Statistics, available from (https://www.cdc.gov/nchs/data/hestat/obesity_child_15_16/obesity_child_15_16.pf), accessed on September 30, 2022. The data on adult obesity prevalence by the state were collected through CDC’s Behavioral Risk Factor Surveillance System (BRFSS), and downloadable from the CDC website (https://www.cdc.gov/obesity/data/prevalence-maps.html#downloads). The data on AC ownership was collected by Annual Characteristics of New Housing Surveys conducted by the US Census Bureau and funded by the Department of Housing and Urban Development. It is available from (https://www.census.gov/construction/chars/).

## Notes

### Competing Interest Statement

The authors have declared no competing interest.

### Funding Statement

This study did no received any funding

## References

1. Mozaffarian, D., Perspective: Obesity-an unexplained epidemic. Am. J. Clin. Nutr. 2022, 115, (6), 1445–1450.

2. McAllister, E. J.; Dhurandhar, N. V.; Keith, S. W.; Aronne, L. J.; Barger, J.; Baskin, M.; Benca, R. M.; Biggio, J.; Boggiano, M. M.; Eisenmann, J. C.; Elobeid, M.; Fontaine, K. R.; Gluckman, P.; Hanlon, E. C.; Katzmarzyk, P.; Pietrobelli, A.; Redden, D. T.; Ruden, D. M.; Wang, C.; Waterland, R. A.; Wright, S. M.; Allison, D. B., Ten putative contributors to the obesity epidemic. Crit. Rev. Food Sci. Nutr. 2009, 49, (10), 868–913.

3. Frieden, T. R.; Ethier, K.; Schuchat, A., Improving the Health of the United States With a “Winnable Battles” Initiative. JAMA 2017, 317, (9), 903–904.

4. Zylke, J. W.; Bauchner, H., The Unrelenting Challenge of Obesity. JAMA 2016, 315, (21), 2277–8.

5. Johnson, F.; Mavrogianni, A.; Ucci, M.; Vidal-Puig, A.; Wardle, J., Could increased time spent in a thermal comfort zone contribute to population increases in obesity? Obes. Rev. 2011, 12, (7), 543–51.

6. Ludwig, D. S.; Aronne, L. J.; Astrup, A.; de Cabo, R.; Cantley, L. C.; Friedman, M. I.; Heymsfield, S. B.; Johnson, J. D.; King, J. C.; Krauss, R. M.; Lieberman, D. E.; Taubes, G.; Volek, J. S.; Westman, E. C.; Willett, W. C.; Yancy, W. S.; Ebbeling, C. B., The carbohydrate-insulin model: a physiological perspective on the obesity pandemic. Am. J. Clin. Nutr. 2021, 114, (6), 1873–1885.

7. Moellering, D. R.; Smith, D. L., Jr., Ambient Temperature and Obesity. Curr Obes Rep 2012, 1, (1), 26–34.

8. Bradtmueller, J. P.; Foley, S. P. In Historical trends of exterior wall materials used in us residential construction, 50th ASC annual international conference proceedings, 2014; 2014.

9. Thompson, K.; Rekowski, A.; Imel, L., Redesign of the Survey of Construction Permit Sample Estimation Procedure. MCD Workino Paper Number: Census/MCD/WP-99/xx, US Census Bureau 1999.

10. National Center for Health Statistics, C. f. D. C. a. P. National Health and Nutrition Examination Survey - About the National Health and Nutrition Examination Survey; National Center for Health Statistics, Centers for Disease Control and Prevention: 2009.

11. Fryar, C. D.; Carroll, M. D.; Ogden, C. L., Prevalence of overweight, obesity, and severe obesity among children and adolescents aged 2–19 years: United States, 1963–1965 through 2015–2016. 2018.

12. Ellison-Barnes, A.; Johnson, S.; Gudzune, K., Trends in Obesity Prevalence Among Adults Aged 18 Through 25 Years, 1976-2018. JAMA 2021, 326, (20), 2073–2074.

13. Remington, P. L.; Smith, M. Y.; Williamson, D. F.; Anda, R. F.; Gentry, E. M.; Hogelin, G. C., Design, characteristics, and usefulness of state-based behavioral risk factor surveillance: 1981-87. Public Health Rep. 1988, 103, (4), 366–75.

14. National Center for Health Statistics, Plan and Operation of the Third National Health and Nutrition Examination Survey (NHANES III, 1988-94). In US Department of Health and Human Service, Public Health Service, Center for Disease Control and Prevention: Hyattsville, MD, 1996.

15. Ford, E. S.; Dietz, W. H., Trends in energy intake among adults in the United States: findings from NHANES. Am. J. Clin. Nutr. 2013, 97, (4), 848–53.

16. Kim, H. J.; Chen, H. S.; Byrne, J.; Wheeler, B.; Feuer, E. J., Twenty years since Joinpoint 1.0: Two major enhancements, their justification, and impact. Stat. Med. 2022.

17. Lipsitz, S. R.; Leong, T.; Ibrahim, J.; Lipshultz, S., A partial correlation coefficient and coefficient of determination for multi-variate normal repeated measures data. Journal of the Royal Statistical Society: Series D (The Statistician) 2001, 50, (1), 87–95.

18. Kingma, B. R.; Frijns, A. J.; Schellen, L.; van Marken Lichtenbelt, W. D., Beyond the classic thermoneutral zone: Including thermal comfort. Temperature (Austin) 2014, 1, (2), 142–9.

19. Cannon, B.; Nedergaard, J., Brown adipose tissue: function and physiological significance. Physiol. Rev. 2004, 84, (1), 277–359.

20. Chaffee, R. R.; Roberts, J. C., Temperature acclimation in birds and mammals. Annu. Rev. Physiol. 1971, 33, 155–202.

21. Herrington, L., The heat regulation of small laboratory animals at various environmental temperatures. American Journal of Physiology-Legacy Content 1940, 129, (1), 123–139.

22. Davis, T. R., THE INFLUENCE OF CLIMATE ON NUTRITIONAL REQUIREMENTS. Am. J. Public Health Nations Health 1964, 54, (12), 2051–67.

23. Erikson, H.; Krog, J.; Andersen, K. L.; Scholander, P. F., The critical temperature in naked man. Acta Physiol. Scand. 1956, 37, (1), 35–9.

24. Deem, J. D.; Muta, K.; Ogimoto, K.; Nelson, J. T.; Velasco, K. R.; Kaiyala, K. J.; Morton, G. J., Leptin regulation of core body temperature involves mechanisms independent of the thyroid axis. Am. J. Physiol. Endocrinol. Metab. 2018, 315, (4), E552–e564.

25. Kaiyala, K. J.; Ogimoto, K.; Nelson, J. T.; Schwartz, M. W.; Morton, G. J., Leptin signaling is required for adaptive changes in food intake, but not energy expenditure, in response to different thermal conditions. PLoS One 2015, 10, (3), e0119391.

26. Yu, S.; Qualls-Creekmore, E.; Rezai-Zadeh, K.; Jiang, Y.; Berthoud, H. R.; Morrison, C. D.; Derbenev, A. V.; Zsombok, A.; Münzberg, H., Glutamatergic Preoptic Area Neurons That Express Leptin Receptors Drive Temperature-Dependent Body Weight Homeostasis. J. Neurosci. 2016, 36, (18), 5034–46.

27. Mader, T. L., Environmental stress in confined beef cattle. J. Anim. Sci. 2003, 81, (14_suppl_2), E110–E119.

28. Collin, A.; van Milgen, J.; Dubois, S.; Noblet, J., Effect of high temperature on feeding behaviour and heat production in group-housed young pigs. Br. J. Nutr. 2001, 86, (1), 63–70.

29. Yahav, S.; Straschnow, A.; Plavnik, I.; Hurwitz, S., Effects of diurnally cycling versus constant temperatures on chicken growth and food intake. Br. Poult. Sci. 1996, 37, (1), 43–54.

30. van Marken Lichtenbelt, W. D.; Vanhommerig, J. W.; Smulders, N. M.; Drossaerts, J. M.; Kemerink, G. J.; Bouvy, N. D.; Schrauwen, P.; Teule, G. J., Cold-activated brown adipose tissue in healthy men. N. Engl. J. Med. 2009, 360, (15), 1500–8.

31. Buemann, B.; Astrup, A.; Christensen, N. J.; Madsen, J., Effect of moderate cold exposure on 24-h energy expenditure: similar response in postobese and nonobese women. Am. J. Physiol. 1992, 263, (6), E1040–5.

32. Westerterp-Plantenga, M. S.; van Marken Lichtenbelt, W. D.; Cilissen, C.; Top, S., Energy metabolism in women during short exposure to the thermoneutral zone. Physiol. Behav. 2002, 75, (1-2), 227–35.

33. Dauncey, M. J., Influence of mild cold on 24 h energy expenditure, resting metabolism and diet-induced thermogenesis. Br. J. Nutr. 1981, 45, (2), 257–67.

34. van Marken Lichtenbelt, W. D.; Westerterp-Plantenga, M. S.; van Hoydonck, P., Individual variation in the relation between body temperature and energy expenditure in response to elevated ambient temperature. Physiol. Behav. 2001, 73, (1-2), 235–42.

35. van Marken Lichtenbelt, W.; Hanssen, M.; Pallubinsky, H.; Kingma, B.; Schellen, L., Healthy excursions outside the thermal comfort zone. Building Research & Information 2017, 45, (7), 819–827.

36. Hanssen, M. J.; Hoeks, J.; Brans, B.; van der Lans, A. A.; Schaart, G.; van den Driessche, J. J.; Jörgensen, J. A.; Boekschoten, M. V.; Hesselink, M. K.; Havekes, B.; Kersten, S.; Mottaghy, F. M.; van Marken Lichtenbelt, W. D.; Schrauwen, P., Short-term cold acclimation improves insulin sensitivity in patients with type 2 diabetes mellitus. Nat. Med. 2015, 21, (8), 863–5.

37. Pallubinsky, H.; Schellen, L.; Kingma, B. R. M.; Dautzenberg, B.; van Baak, M. A.; van Marken Lichtenbelt, W. D., Thermo-physiological adaptations to passive mild heat acclimation. Temperature (Austin) 2017, 4, (2), 176–186.

38. Pallubinsky, H.; Phielix, E.; Dautzenberg, B.; Schaart, G.; Connell, N. J.; de Wit-Verheggen, V.; Havekes, B.; van Baak, M. A.; Schrauwen, P.; van Marken Lichtenbelt, W. D., Passive exposure to heat improves glucose metabolism in overweight humans. Acta Physiol. (Oxf.) 2020, 229, (4), e13488.

39. Stettler, N., Nature and strength of epidemiological evidence for origins of childhood and adulthood obesity in the first year of life. Int. J Obes (Lond.) 2007, 31, (7), 1035–43.

40. Singh, A. S.; Mulder, C.; Twisk, J. W.; van Mechelen, W.; Chinapaw, M. J., Tracking of childhood overweight into adulthood: a systematic review of the literature. Obes. Rev. 2008, 9, (5), 474–88.

41. Starc, G.; Strel, J., Tracking excess weight and obesity from childhood to young adulthood: a 12-year prospective cohort study in Slovenia. Public Health Nutr. 2011, 14, (1), 49–55.

42. Glass, L.; Silverman, W. A.; Sinclair, J. C., Effect of the thermal environment on cold resistance and growth of small infants after the first week of life. Pediatrics 1968, 41, (6), 1033–46.

43. Snook, K. R.; Hansen, A. R.; Duke, C. H.; Finch, K. C.; Hackney, A. A.; Zhang, J., Change in Percentages of Adults With Over-weight or Obesity Trying to Lose Weight, 1988-2014. JAMA 2017, 317, (9), 971–973.

44. Centers for Disease Control and Preventions, U., Behavioral Risk Factor Surveillance System. http://www.cdc.gov/nccdphp/publications/aag/brfss.htm 2013.

45. Yun, S.; Zhu, B. P.; Black, W.; Brownson, R. C., A comparison of national estimates of obesity prevalence from the behavioral risk factor surveillance system and the National Health and Nutrition Examination Survey. Int. J Obes (Lond.) 2006, 30, (1), 164–70.

46. Mokdad, A. H.; Bowman, B. A.; Ford, E. S.; Vinicor, F.; Marks, J. S.; Koplan, J. P., The continuing epidemics of obesity and diabetes in the United States. JAMA 2001, 286, (10), 1195–200.

47. Birol, F., The future of cooling: opportunities for energy-efficient air conditioning. International Energy Agency 2018.

48. Syndicus, M.; Wiese, B. S.; van Treeck, C., Too hot to carry on? Disinclination to persist at a task in a warm office environment. Ergonomics 2018, 61, (4), 476–481.

49. Maula, H.; Hongisto, V.; Östman, L.; Haapakangas, A.; Koskela, H.; Hyönä, J., The effect of slightly warm temperature on work performance and comfort in open-plan offices - a laboratory study. Indoor Air 2016, 26, (2), 286–97.

50. He, F.; Wei, J.; Dong, Y.; Liu, C.; Zhao, K.; Peng, W.; Lu, Z.; Zhang, B.; Xue, F.; Guo, X.; Jia, X., Associations of ambient temperature with mortality for ischemic and hemorrhagic stroke and the modification effects of greenness in Shandong Province, China. Sci. Total Environ. 2022, 851, (Pt 1), 158046.

51. Gasparrini, A.; Guo, Y.; Hashizume, M.; Lavigne, E.; Zanobetti, A.; Schwartz, J.; Tobias, A.; Tong, S.; Rocklöv, J.; Forsberg, B.; Leone, M.; De Sario, M.; Bell, M. L.; Guo, Y. L.; Wu, C. F.; Kan, H.; Yi, S. M.; de Sousa Zanotti Stagliorio Coelho, M.; Saldiva, P. H.; Honda, Y.; Kim, H.; Armstrong, B., Mortality risk attributable to high and low ambient temperature: a multicountry observational study. Lancet 2015, 386, (9991), 369–75.

52. Dietz, W. H.; Pryor, S., How Can We Act to Mitigate the Global Syndemic of Obesity, Undernutrition, and Climate Change? Curr Obes Rep 2022, 11, (3), 61–69.

