## Supplemental Figure 1 for "Spatiotemporal Correlation between Obesity Prevalence and the Percentage of Households using Air-conditioners in the United States"

Supplemental Figure 1: United States Census Regions and Divisions

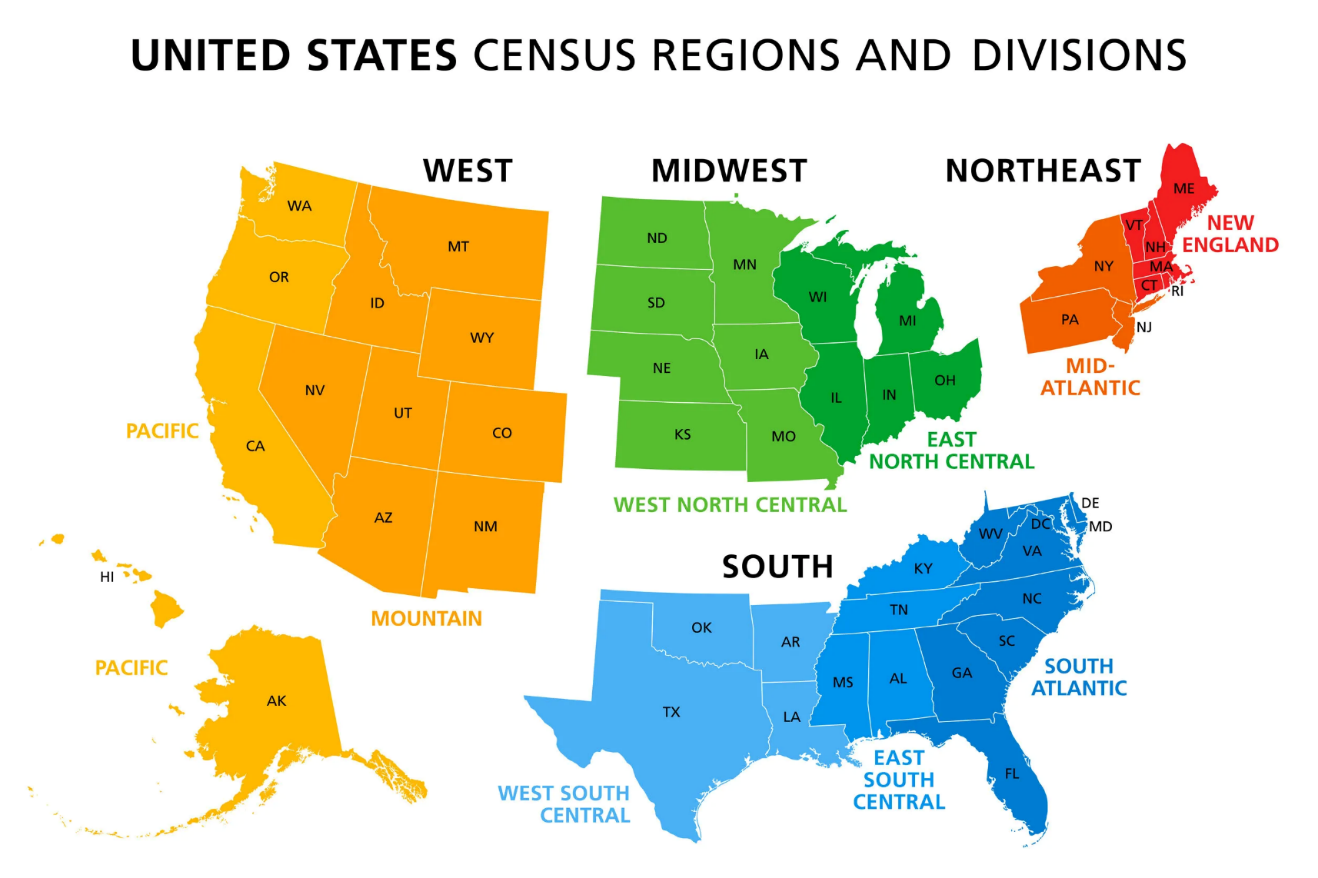

Supplemental Figure 1: United States Census Regions and Divisions

1. Source: U.S. Census Bureau. It is not protected by the U.S. Copyright Act and can be used without obtaining permission or paying a copyright fee. Available from <https://www.census.gov/construction/chars/definitions/#r> accessed on Sept 10, 2022.
2. The Northeast region includes Connecticut, Maine, Massachusetts, New Hampshire, New Jersey, New York, Pennsylvania, Rhode Island, and Vermont; the Midwest region includes Illinois, Indiana, Iowa, Kansas, Michigan, Minnesota, Missouri, Nebraska, North Dakota, Ohio, South Dakota, and Wisconsin; South region includes Alabama, Arkansas, Delaware, District of Columbia (excluded from current analyses due to missing obesity data), Florida, Georgia, Kentucky, Louisiana, Maryland, Mississippi, North Carolina, Oklahoma, South Carolina, Tennessee, Texas, Virginia, and West Virginia; and the West region includes Alaska, Arizona, California, Colorado, Hawaii, Idaho, Montana, Nevada, New Mexico, Oregon, Utah, Washington, and Wyoming.

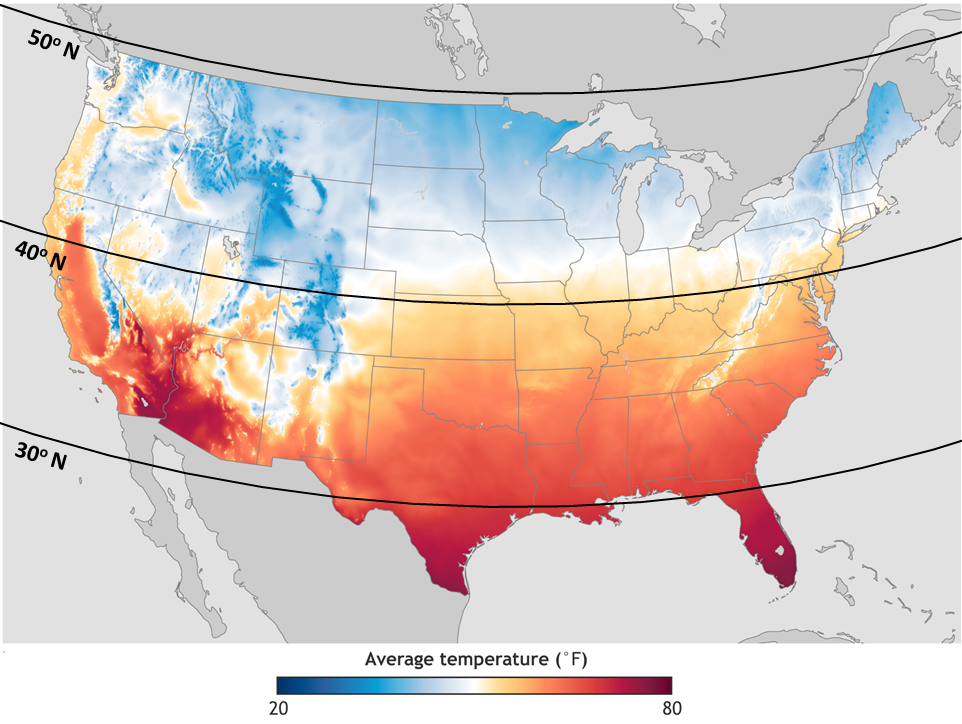

Supplemental Figure 2: U.S. Annual Average Temperature across Regions, The contiguous United States. 1991 - 2020

Source: National Oceanic and Atmospheric Administration. It is not protected by the U.S. Copyright Act and can be used without obtaining permission or paying a copyright fee. Available from <https://www.climate.gov/news-features/featured-images/new-maps-annual-average-temperature-and-precipitation-us-climate>. Accessed on Sept 10, 2022

Supplemental Table 1. Correlation between Obesity Prevalence and Ownership of AC, Dietary intake 1977 – 2016, United States ^1,2,3^

|  | | Pearson correlation | | | Spearman correlation | | Adjusted partial correlation | |
| --- | --- | --- | --- | --- | --- | --- | --- | --- |
| Age groups |  | Ownership of air conditioners | | Dietary intake | Ownership of air conditioners | Dietary intake | Ownership of air conditioners | Dietary intake |
| Children 2 to 5 years | coefficient | | 0.614 | 0.567 | 0.370 | 0.351 | 0.008 | 0.000007 |
|  | *p-value* | | 0.263 | 0.290 | 0.263 | 0.290 | 0.207 | 0.388 |
| Children 6 to 11 years | coefficient | | 0.804 | 0.690 | 0.374 | 0.023 | 0.027 | 0.000013 |
|  | *p-value* | | 0.257 | 0.947 | 0.257 | 0.947 | 0.026 | 0.263 |
| Children 12 to 19 years | coefficient | | 0.943 | 0.674 | 0.858 | -.137 | 0.046 | 0.000001 |
|  | *p-value* | | 0.001 | 0.689 | 0.001 | 0.689 | <0.001 | 0.584 |
| Adult 18-25 years | coefficient | | 0.916 | 0.790 | 0.774 | 0.327 | 0.046 | 0.000002 |
|  | *p-value* | | 0.005 | 0.326 | 0.005 | 0.326 | <0.000 | 0.584 |

1. The dietary energy intakes were estimated by Ford and Dietz ^15^ and Mozaffarian^1^ using data from National Health and Nutrition Examination Surveys.
2. The estimations were made by Fryar et al, ^11^ Prevalence of Overweight, Obesity, and Severe Obesity Among Children and Adolescents Aged 2–19 Years: the United States, 1963–1965 Through 2015–2016. Health E-State, September 2018, National Center for Health Statistics.
3. The data on AC ownership was collected by Annual Characteristics of New Housing Surveys conducted by the US Census Bureau and funded by the Department of Housing and Urban Development. It is available from <https://www.census.gov/construction/chars/>).
